# The risk of severe COVID-19: hospital and ICU admission rates in Norway

**DOI:** 10.1101/2020.07.16.20155358

**Authors:** Morten Munkvik, Ingvild Vatten Alsnes, Lars Vatten

## Abstract

**Background:** Epidemiological studies of COVID-19 with population based information may add to the knowledge needed to prioritise resources and advice on how restrictive measures should be targeted. This study provides admission rates to hospitals and intensive care units (ICU) in Norway, aiming to better understand the risk of severe COVID-19 infection.

**Methods:** Data from official reports from The Norwegian Institute of Public Health (NIPH) and the Norwegian Directorate of Health were used to calculate admission rates to hospitals and to ICU per 100 000 inhabitants. We compared rates of hospitalisation between the four health regions and provide separate rates for Oslo. We also assessed national admissions to ICU stratified by age.

**Results:** The admission rate in the south-eastern region was 3.1 per 100 000, and the rate for Oslo was 5.8. Compared to the western region (reference), the Oslo rate was 4.0 times (confidence interval (CI) 3.0-5.5) higher. In Norway as a whole, the rate of ICU admissions was 3.9 per 100 000, and in the age groups 60-69 and 70-79, ICU rates were 10.3 and 11.5, respectively. These rates were 9.5 (CI 6.3-14.3) and 10.6 (CI 6.9-16.2) times higher compared to people younger than 50 years.

**Conclusion:** Hospital admissions due to Covid-19 are much higher in Oslo than anywhere else in Norway, and in the country as a whole, ICU admissions are highest among people 60-79 years of age. These results and more detailed data could provide better advice on how restrictions can be safely lessened.

## Introduction

The general knowledge about COVID-19 has gone from almost nil to being relatively high in a few months. Nonetheless, more extensive knowledge is needed to diagnose and treat the infection correctly. Much knowledge is based on prediction models that include information from different countries. So far, risk factors for Covid-19 in other countries seem to apply also in Norway. Still, we need epidemiological descriptions of the COVID-19 outbreak on a national level. This could be useful in deciding who precisely should be protected from the virus, how to prioritise the use of resources, and which restrictive measures are particularly useful to prevent spreading of the virus. Thus, population based information may teach us that certain parts of society can manage with less harsh restrictions than others, and that improved targeting of measures may reduce the negative socio-economic impact.

As of April 27^th^, 2020, there were 3,043,800 confirmed cases of COVID-19 in the world, and 214,489 deaths.(1) The complete death toll will not be clear for a long time. Limited test capacity obscures the true incidence of infected cases, because large scale representative testing is typically not performed. Data from China suggest that one in five COVID-19 patients has very mild symptoms, and these patients would not be subjected to testing in most countries.(2) In Norway, only symptomatic cases are tested, and yet 95-99 % are test negative, indicating that the prevalence of COVID-19 is low compared to other airway infections.(3) Nevertheless, undetected asymptomatic cases of COVID-19 may infect others, who might develop severe disease.(4) Whereas the number of confirmed cases does not reliably reflect its incidence, hospital admissions and mortality rates of COVID-19 are good indicators of severe COVID-19 in Norway.

Patients with COVID-19 may be divided into four categories: patients with mild symptoms who are isolated at home; patients with more severe symptoms who are admitted to a local care facility; patients with severe symptoms who are hospitalized; or patients with severe symptoms who require intensive care. The three latter categories are at elevated risk of severe illness and death.

Here we report the rate of admissions to hospitals in the four health regions of Norway as of April 27^th^. We also report admission rates for the population of Oslo, the capital of Norway. In addition, we report cumulative admission rates to intensive care units (ICU) in Norway from the first confirmed case (February 26^th^) until April 27^th^ and compare the need for intensive care between different age groups.

## Methods

The Norwegian Institute of Public Health (NIPH) and the Norwegian Directorate of Health (Hdir) publish daily reports related to COVID-19 in Norway.(5, 6) For Intensive Care Units (ICU), the admission rates are cumulative, as consecutively recorded in the national registry of ICU patients. All data was part of public reports, thus no ethical approval was necessary. We studied rates of admission to ICU for the whole country and stratified the admissions according to age groups in the population.

In contrast to the cumulative ICU data, the total number of patients admitted to hospital because of COVID-19 is not cumulatively presented in the daily reports from the health authorities. Therefore, we used data from the most recent report (April 27^th^ 2020) in the analyses of hospital admissions. However, by following the daily reports of admissions prior to that date, we could also analyze daily admission rates over time, and present them graphically.

We calculated admission rates due to COVID-19 per 100 000 inhabitants, applying the most recent population numbers, including age distribution, provided by Statistics Norway as of January 1^st^ 2020. These admission rates reflect the incidence of severe COVID-19 infection. We compared the rates between the four health regions in Norway; the south-eastern region, the western region, the region of the middle part of the country, and the northern region. Oslo, the capital of Norway, belongs to the south-eastern health region, but we also present separate admission rates for Oslo. In the comparisons, we used the admission rate of the western region as the reference, and present rate ratios of admissions between regions with 95% confidence intervals. The statistical analyses were conducted with the statistical program R version 3.6.3.

## Results

Table 1 shows that on April 27^th^, 94 COVID-19 patients were admitted to hospitals in health region South-East, yielding a rate of 3.10 per 100 000. The separate rate for Oslo was 5.77, which is 86% higher compared to the rest of that health region. And compared to the western health region, the admission rate in Oslo was 4.03 (confidence interval (CI) 2.96 – 5.49) times higher.

**Tabell 1.** Hospitalized covid-19 patients on April 27^th^ 2020.

| Region | Population<sup>1</sup> | Hospitalized<sup>2</sup> | Rate<sup>3</sup> | Rate ratio | 95% confidence interval |
| --- | --- | --- | --- | --- | --- |
| Western | 1 117 586 | 16 | 1.43 | 1.0 (Ref) |  |
| South-eastern | 3 032 670 | 94 | 3.10 | 2.17 | 1.77 – 2.65 |
| Central | 732 777 | 7 | 0.96 | 0.67 | 0.32 – 1.40 |
| Northern | 486 975 | 8 | 1.64 | 1.61 | 0.57 – 2.29 |
| Total | 5 370 008 | 125 | 2.33 |  |  |
| Oslo | 693 491 | 40 | 5.77 | 4.03 | 2.96 – 5.49 |
| South-eastern<sup>4</sup> | 2 339 179 | 54 | 2.31 | 1.61 | 1.23 – 2.11 |

<sup>1</sup>Updated January 1st 2020
<sup>2</sup>On April 27th 2020 (not cumulative number)
<sup>3</sup>Per 100.000
<sup>4</sup>South-eastern region excluding Oslo

Figure 1 illustrates how the rate ratios of hospital admissions across health regions have developed over time. Compared to the rates in the western region, the rate ratios on April 5^th^ (2.50, 0.96, 1.05, and 4.01, for the south-western region (excluding Oslo), middle of Norway, northern Norway and Oslo, respectively) were roughly similar to the rate ratios on April 27^th^ (2.17, 0.67, 1.15 and 4.03). The figure also shows that the number of COVID-19 patients admitted to hospital has decreased during this time period.

**Figure 1.**
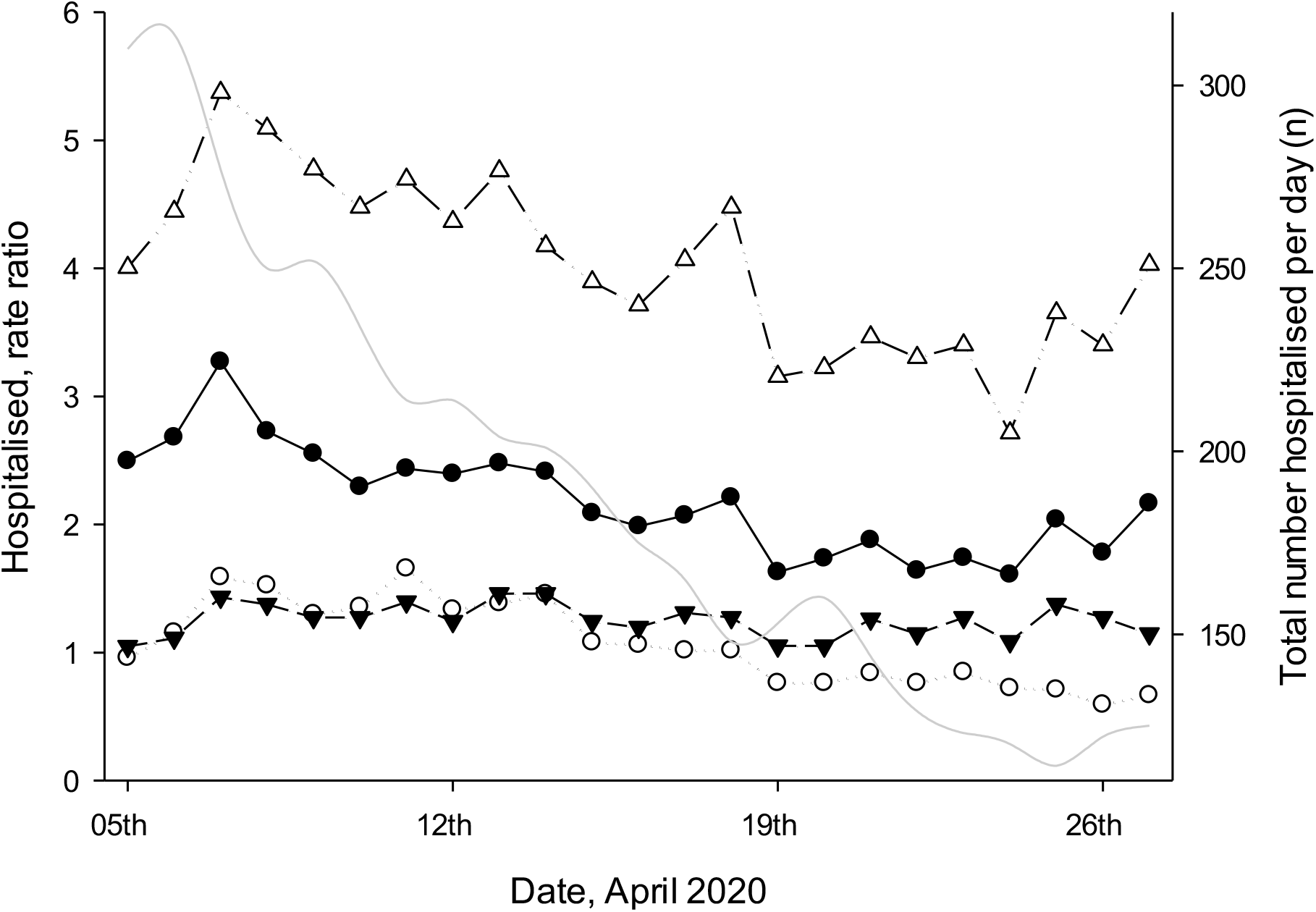
Rate ratios of hospital admissions across health regions relative to the western region of Norway, and total number of hospitalized patients. South-eastern region of Norway (filled circle), middle region (open circle), northern region (filled triangle) and the city of Oslo (open triangle). Gray line is total number of hospitalised patients.

Table 2 includes all COVID-19 patients who were consecutively admitted to intensive care units (ICU) in the country. The total rate is 3.93 per 100 000, but in the 60-69 year age group, the rate of admission is 10.29 per 100 000, and among people 70-79 years of age, the corresponding rate is 11.47. Using the rate of ICU admission among people younger than 50 years as the reference, we found that the admission rate in the 60-69 year age group was 9.50 (CI 6.31 – 14.31) higher, and correspondingly,10.59 (CI 6.92 – 16.20) times higher among those 70-79 years of age. Compared to the reference, the rate of admission to ICU was 5.20 (CI 2.76 – 9.78) higher among people 80 years or older.

**Tabel 2.** Cumulative number ICU patients due to covid-19 as of April 27^th^ 2020, per age group.

| Age | Population<sup>1</sup> | ICU patients<sup>2</sup> | Rate<sup>3</sup> | Rate ratio | 95% confidence interval |
| --- | --- | --- | --- | --- | --- |
| <50 | 3 414 711 | 37 | 1.08 | 1.0 (Ref) |  |
| 50-59 | 703 830 | 51 | 7.25 | 6.69 | 4.38 – 10.21 |
| 60-69 | 582 995 | 60 | 10.29 | 9.50 | 6.31 – 14.31 |
| 70-79 | 435 834 | 50 | 11.47 | 10.59 | 6.92 – 16.20 |
| ≥80 | 230 710 | 13 | 5.63 | 5.20 | 2.76 – 9.78 |
| Total | 5 368 080 | 211 | 3.93 |  |  |

<sup>1</sup>Populasjon as of January 1<sup>st</sup> 2020.
<sup>2</sup>Cumulative ICU patients due to covid-19 up to April 27<sup>th</sup> 2020.
<sup>3</sup>Per 100.000

## Discussion

As of April 27^th^, the likelihood for hospitalisation due to COVID-19 was four times higher in Oslo compared to the western health region of Norway. The admission rate for the other health regions was also substantially lower than in Oslo. In the country as a whole, the likelihood of needing ICU treatment was about ten times higher for people between 60 to79 years of age compared to people below the age of 50 years. The rate of ICU treatment rises sharply after the age of 50, and for those between 50-59 the risk is almost 7 times higher compared to those below 50. For people 80 years or older, the corresponding rate is 5 times higher, most likely reflecting that elderly patients are less likely to be treated in intensive cure units, but more likely to be treated locally (mean age of death from COVID-19 in Norway is 82 years of age).(7) Our most important finding is that Oslo has a much higher rate of hospitalised COVID-19 cases compared to the rest of the country.

We used data that are available from daily reports published by national health authorities. Optimally, complementary information on age, gender, comorbidity and place of residence should also be available, but is missing from those reports. It is difficult to see that more complete information would have interfered with laws of privacy or the duty of confidentiality. Thus, the authorities′ unwillingness to share information with the public is a limitation of this study. Nonetheless, we are not aware of comparable epidemiological studies on hospital and ICU admission rates due to COVID-19. Many international studies are based on prediction models and rely on assumptions that might not be readily applicable to Norway.

The number of admitted patients has now shown a consistent decrease, and the death rate has flattened out, and therefore, it has been argued that the situation in Norway is under control. However, our findings suggest that the situation in Oslo deviates strongly from the rest of the country, and that the capital requires special attention.

As the society is opening up – schools have started to re-open and social distancing restrictions are lessened – the situation in Oslo needs to be monitored very carefully. Oslo is the obvious epi-center of the epidemic in Norway, from where new waves of infection are likely to arise. Therefore, targeted testing and contact tracing should be prioritized in Oslo, whereas in other regions, with modest transmission of SARS-CoV2, strategic testing of health care workers or others with close contacts with individuals at increased risk, may be sufficient. It is especially important to limit infections among people older than 60 years of age, with particular emphasis on shielding those 80 years or older.

We would need more detailed data than what is presently available in order to provide reliable advice that may justify further reductions of the restrictions. It is a paradox that better ways to meet the pandemic could lie hidden in these undisclosed data. Apart from being the epi-center of the COVID-19 pandemic in Norway, Oslo is also the place in the country where the infection is likely to flare up again.

## Data Availability

All data was part of public reports, thus no ethical approval was necessary.

